# Preventing respiratory illness in older adults aged 60 years and above living in long-term care

**DOI:** 10.1101/2020.03.19.20039081

**Authors:** Patricia Rios, Amruta Radhakrishnan, Sonia M. Thomas, Nazia Darvesh, Sharon E. Straus, Andrea C. Tricco

**Affiliations:** Li Ka Shing Knowledge Institute, St. Michael's Hospital, Unity Health Toronto

## Abstract

**Background:** The overall objective of this rapid overview of reviews (overview hereafter) was to identify evidence from systematic reviews (SRs) for infection control and prevention practices for adults aged 60 years and older in long-term care settings.

**Methods:** Comprehensive searches in MEDLINE, EMBASE, the Cochrane Library, biorxiv.org/medrxiv.org, clinicaltrials.gov and the Global Infectious Disease Epidemiology Network (GIDEON) were carried out in early March 2020. Title/abstract and full-text screening, data abstraction, and quality appraisal (AMSTAR 2) were carried out by single reviewers.

**Results:** A total of 6 SRs published between 1999 and 2018 were identified and included in the overview. The SRs included between 1 and 37 primary studies representing between 140 to 908 patients. All of the primary studies included in the SRs were carried out in long-term care facilities (LTCF) and examined pharmacological, non-pharmacological, or combined interventions. One high quality SR found mixed results for the effectiveness of hand hygiene to prevent infection (2 studies statistically significant positive results, 1 study non-statistically significant results). One moderate quality SR with meta-analysis found a moderate non-statistically significant effect for personal protective equipment (PPE) in preventing infection and found no statistically significant results for the effectiveness of social isolation. One moderate quality SR reported statically significant evidence for the effectiveness of amantadine and amantadine + PPE to prevent infection with respiratory illness in LTCF.

**Conclusion:** The current evidence suggests that with antiviral chemoprophylaxis with adamantine is effective in managing respiratory illness in residents of long-term care facilities. The rest of the strategies can be used in long-term care facilities, yet have limited evidence supporting their use from systematic reviews.

## INTRODUCTION

### Purpose and Research Questions

The Infection Prevention & Control of the World Health Organization (WHO) Health Emergency Programme presented a query on preventing and managing COVID-19 in older adults aged 60 years and above living in long-term care facilities including privately paid for and publicly paid for settings with a 5-business day timeline. According to the WHO, "long-term care covers those activities undertaken by others to ensure that people with, or at risk of, a significant ongoing loss of intrinsic capacity can maintain a level of functional ability consistent with their basic rights, fundamental freedoms and human dignity” (https://www.who.int/ageing/long-term-care/WHO-LTC-series-subsaharan-africa.pdf?ua=1). Examples of long-term care include nursing homes, charitable homes, municipal homes, long-term care hospitals, long-term care facilities, skilled nursing facilities, convalescent homes, and assisted living facilities (https://www.canada.ca/en/health-canada/services/home-continuing-care/long-term-facilities-based-care.html).

The overall objective of this rapid overview of reviews (overview hereafter) was to identify evidence on infection protection and control measures for adults aged 60 years and older in long-term care settings from systematic reviews. In order to focus the research question to increase feasibility, we proposed the following key research questions:

1. What are the infection prevention and control practices/measures for preventing or reducing respiratory viruses (including coronavirus and influenza) in older adults aged 60 years and above living in long-term care?
2. How do infection prevention and control practices differ for adults aged 60 years and above living in long-term care with respiratory illness and severe comorbidities or frailty differ than those without such severe comorbidities/frailty?
3. How do infection prevention and control practices differ for adults aged 60 years and above living in long-term care with respiratory illness from low- and middle-income economy countries (LMIC) differ than those living in high-income economy countries and do differences exist across different cultural contexts?

## METHODS

The rapid overview was guided by the Cochrane Handbook for Systematic Reviews of Interventions^1^ along with the Rapid Review Guide for Health Policy and Systems Research^2^. The team used an integrated knowledge translation approach, with consultation from the knowledge users from the WHO at the following stages: question development, interpretation of results, and draft report. We used the Preferred Reporting Items for Systematic Reviews and Meta-Analyses (PRISMA) Statement^3^ to guide the reporting of our rapid overview results; a PRISMA extension for rapid reviews and a reporting guideline on overviews are currently under development. This rapid overview was completed in conjunction with a rapid review of clinical practice guidelines published in a separate report titled: *Guidelines for preventing respiratory illness in older adults aged 60 years and above living in long-term care: A rapid review of clinical practice guidelines*.

### Protocol

We prepared a brief protocol for this query that is available in Appendix 1. If publication in a peer-reviewed journal is planned in the future, we will register this rapid review with the Open Science Framework (https://osf.io/).

### Literature search

Comprehensive literature searches addressing all research questions were developed by an experienced librarian for MEDLINE, EMBASE, the Cochrane Library, and biorxiv.org/medrxiv.org databases. Grey (i.e., difficult to locate or unpublished) literature was located using keyword searches of relevant terms (e.g. respiratory illness, MERS, coronavirus, SARS, long-term care facilities) in clinicaltrials.gov and GIDEON (Global Infectious Diseases and Epidemiology Network). The full MEDLINE search strategy can be found in Appendix 2. Due to the rapid timelines for this overview, a peer review of the literature search was not conducted.

### Eligibility criteria

The Eligibility criteria followed the PICOST framework and consisted of:

<u>Population:</u> Individuals aged 60 years and above residing in long-term care facilities. The age cut-off for an older adult might be 50 years and above in different LMIC and/or cultural settings. As such, we included these in level 1 screening of titles and abstracts and presented potentially relevant studies in an appendix.

<u>Intervention:</u> Any form of infection control and prevention, such as hand hygiene, respiratory hygiene/etiquette, personal protective equipment (for patients and health care providers), triage (on arrival), source control, isolation, daily monitoring/surveillance for signs and symptoms of respiratory illness (e.g., COVID-19) in residence, environmental cleaning, cleaning of linen and medical equipment used by patients, restrictions on resident movement and transportation, restrictions on visitors, restrictions on travel for health care providers and other long-term care facility staff, waste management, dead body management. Only those measures used to prevent and control respiratory illnesses, including influenza and coronavirus (e.g., COVID-19, MERS, SARS) were included. Interventions focused on preventing bacterial respiratory outbreaks (e.g., strep, pneumonia, klebsiella) or aspiration pneumonia were excluded. Interventions specifically focused on vaccination were excluded, as a vaccine for the coronavirus currently does not exist.

<u>Comparator:</u> One of the interventions listed above or no intervention

<u>Outcomes:</u> Lab-confirmed respiratory illness due to the virus (e.g., SARS, MERS, COVID-19, influenza) [primary outcome], secondary bacterial infection, symptoms, secondary transmission (e.g., other patients, healthcare workers, visitors), goal concordant care, hospitalization, intensive-care unit (ICU) admission, and mortality.

<u>Study designs:</u> We limited inclusion to systematic reviews using the Cochrane definition of a systematic review (https://www.cochrane.org/news/what-are-systematic-reviews).

<u>Time periods:</u> All periods of time and duration of follow-up were eligible.

<u>Other:</u> No other restrictions were imposed.

### Study selection

For both level 1 (title/abstract) and level 2 (full-text) screening, a screening form was prepared based on the eligibility criteria and pilot-tested by the review team using 25 citations prior to level 1 screening and 5 full text articles prior to level 2 screening. Agreement between reviewers was sufficiently high (>75%) in both cases so no further pilot-testing was required. Full screening was completed by a single reviewer for both level 1 and level 2 using Synthesi.SR, the team’s proprietary online software (https://breakthroughkt.ca/login.php).

### Data items and data abstraction

Items for data abstraction included characteristics (e.g., duration of follow-up, study design, country of conduct, multi-center vs. single site, long-term care setting characteristics, such as availability of medical support, characteristics of care staff, family/community engagement, accommodation characteristics, collective practices), patient characteristics (e.g., mean age, age range, co-morbidities), intervention details (e.g., type of intervention, duration and frequency of intervention, timing of intervention), comparator details (e.g., comparator intervention, duration and frequency of intervention, timing of intervention), and outcome results (e.g., lab-confirmed viral respiratory infection, symptoms, secondary transmission, hospitalization, ICU admission, mortality) at the longest duration of follow-up. A standardized data abstraction form was developed. Prior to data abstraction, a calibration exercise was completed to test the form amongst the entire review team using two randomly selected full-text systematic reviews. Following successful completion of the calibration exercise, included reviews were abstracted by single reviewers.

### Risk of bias appraisal

Risk of bias appraisal was carried out by single reviewers using the AMSTAR-2 tool (https://amstar.ca/Amstar-2.php). The AMSTAR-2 tool includes 16 items, plus an overall risk of bias rating that ranges from low risk of bias to high risk of bias, with moderate risk of bias in between. The items focus on methods related to the research question, protocol, literature search, study selection, risk of bias appraisal, data abstraction, meta-analysis, and conflicts of interest.

### Synthesis

Included studies were synthesized descriptively including summary statistics and detailed tables of study characteristics and results. Tables of study results are organized according to study design and where available, information on relevant subgroups were highlighted.

## RESULTS

### Literature Search

The database search returned a total of 3,309 citations, while the grey literature searches returned 42 citations for level 1 screening. A total of 3,225 citations were excluded after level 1 screening. Of the 126 articles screened at level 2, 6 systematic reviews were included (Figure 1).

**Figure 1:**
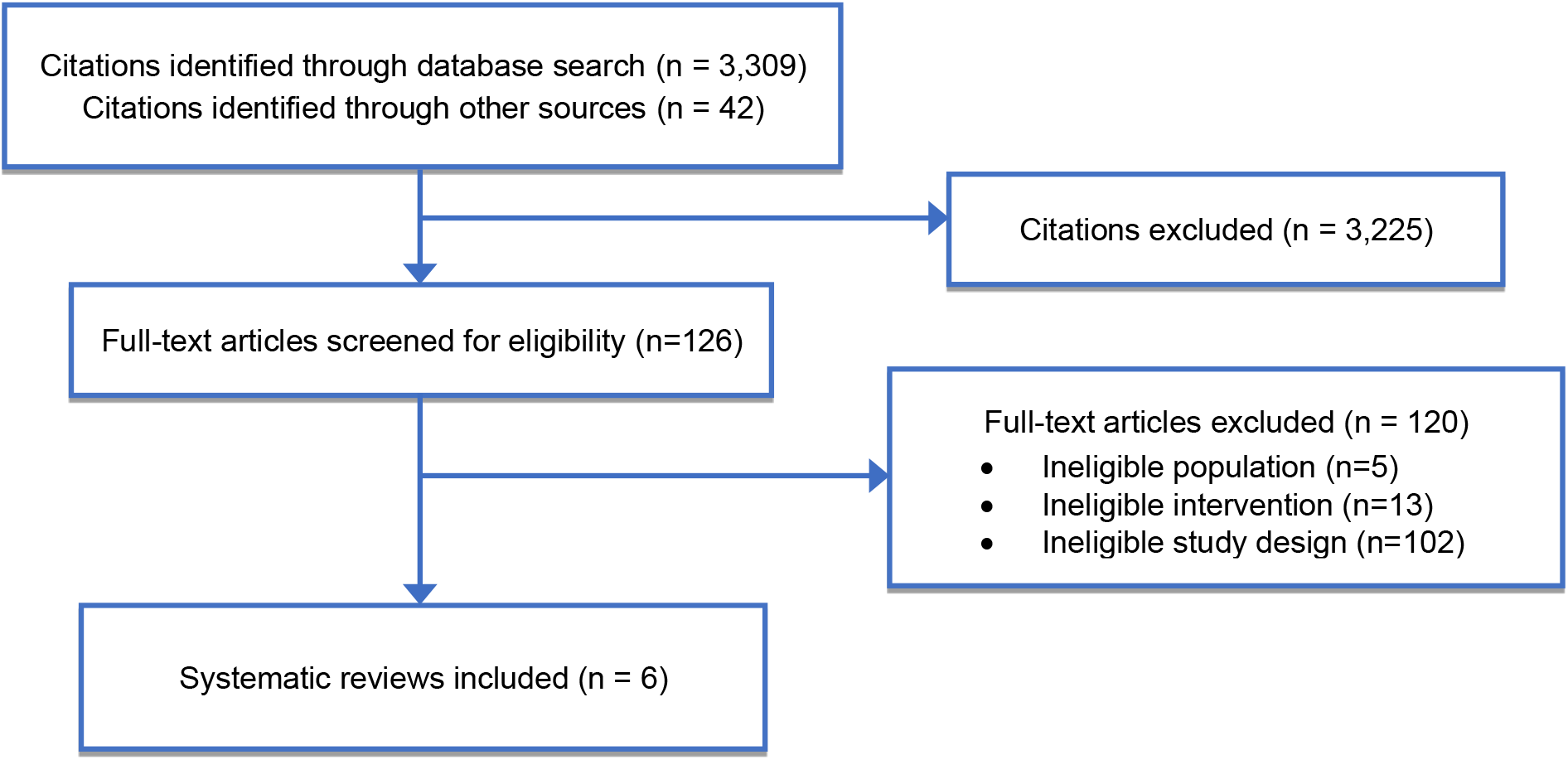
Study flow.

### Characteristics of included systematic reviews

The six systematic reviews were published between 1999 and 2018 (Appendix 3). The number of studies included in the systematic reviews ranged from 1 to 37 (not reported in one systematic review). Only two systematic reviews reported the number of included patients, which ranged from 140 to 908. The systematic reviews included studies that were conducted in long-term care facilities and the interventions were pharmacological (e.g., antiviral chemoprophylaxis), non-pharmacological (e.g., increasing hand hygiene, personal protective equipment, social distancing), and a combination of pharmacological and non-pharmacological.

### Quality Appraisal Results

The six systematic reviews varied in their quality according to the AMSTAR-2 tool, with two being assessed as having a high risk of bias, one with a moderate risk of bias, and three with a low risk of bias (Figure 2, Appendix 4).

**Figure 2:**
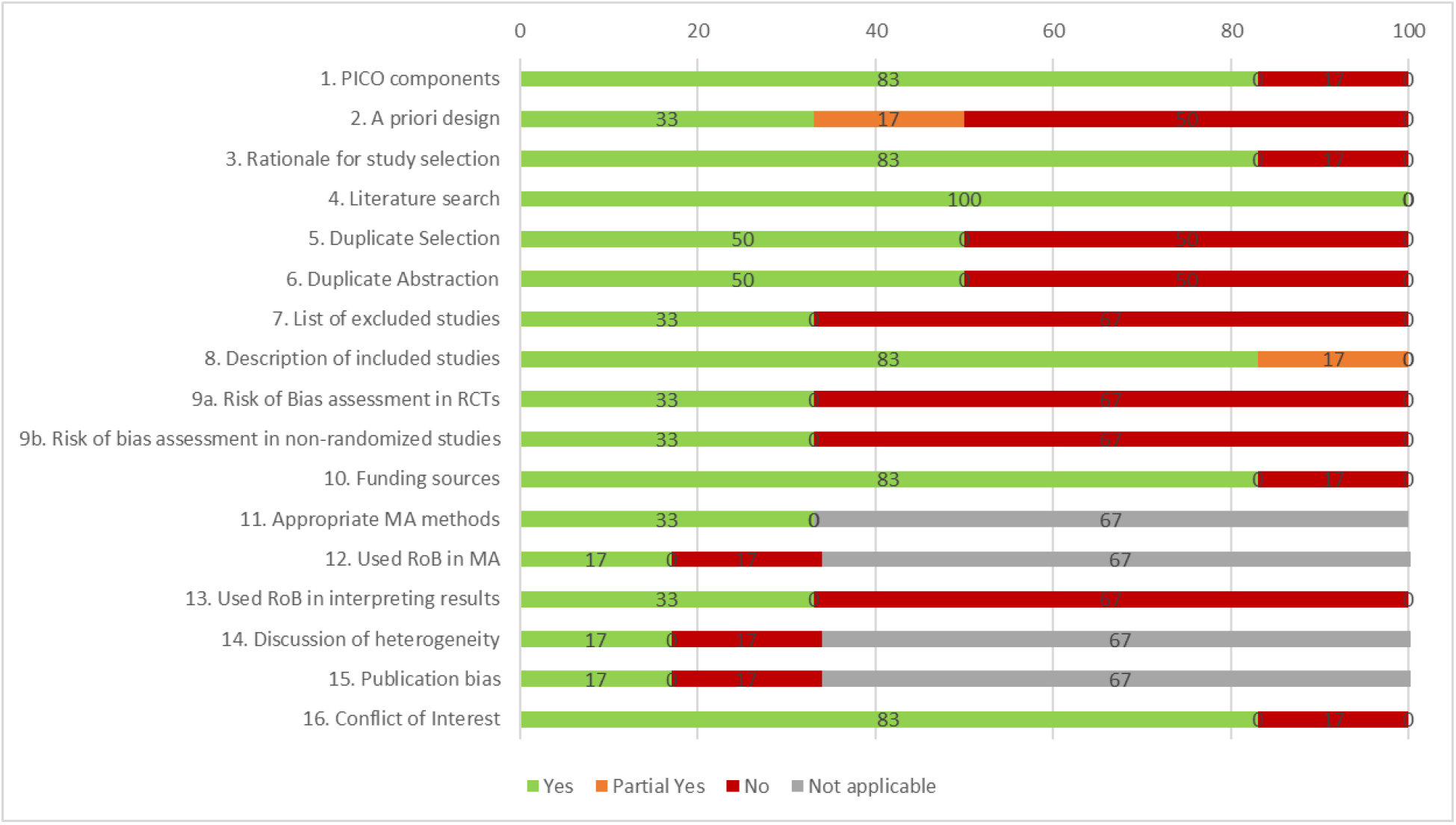
Summary of AMSTAR 2 results.

### Effectiveness Results

#### Preventing respiratory illness in long-term care facilities

One high quality SR found mixed results for the effectiveness of hand hygiene to prevent infection with 2 studies reporting statistically significant positive results in favour of hand hygiene and 1 study reporting non-statistically significant results. One moderate quality SR with meta-analysis found a moderate non-statistically significant effect in favour of personal protective equipment (PPE) in preventing infection. The same SR and meta-analysis also examined the effectiveness of social isolation to prevent infection and found no statistically significant results. One moderate quality SR reported statically significant evidence for the effectiveness of amantadine and amantadine + PPE to prevent the spread of viral respiratory infections in long-term care facilities.

#### Managing respiratory illness in long-term care facilities

Statistically significant results were found from one moderate and one high quality systematic review for the use of amantadine as antiviral chemoprophylaxis for individuals diagnosed with lab-confirmed influenza (Table 1). In addition, statistically significant results were found from one high quality systematic review for the use of amantadine plus personal protective equipment to prevent spread of infection from individuals diagnosed with lab-confirmed influenza. Statistically significant results were not observed in one moderate and one high quality systematic review regarding rimantadine or neuraminidase inhibitors as antiviral chemoprophylaxis. However, statistically significant results were observed in one low quality systematic review for rimantadine as antiviral chemoprophylaxis. Mixed evidence was identified from two low quality systematic reviews for zanamivir as antiviral chemoprophylaxis that reported non-statistically significant reductions in viral respiratory infection rates. The full systematic review results are available in Appendix 5.

**Table 1:**
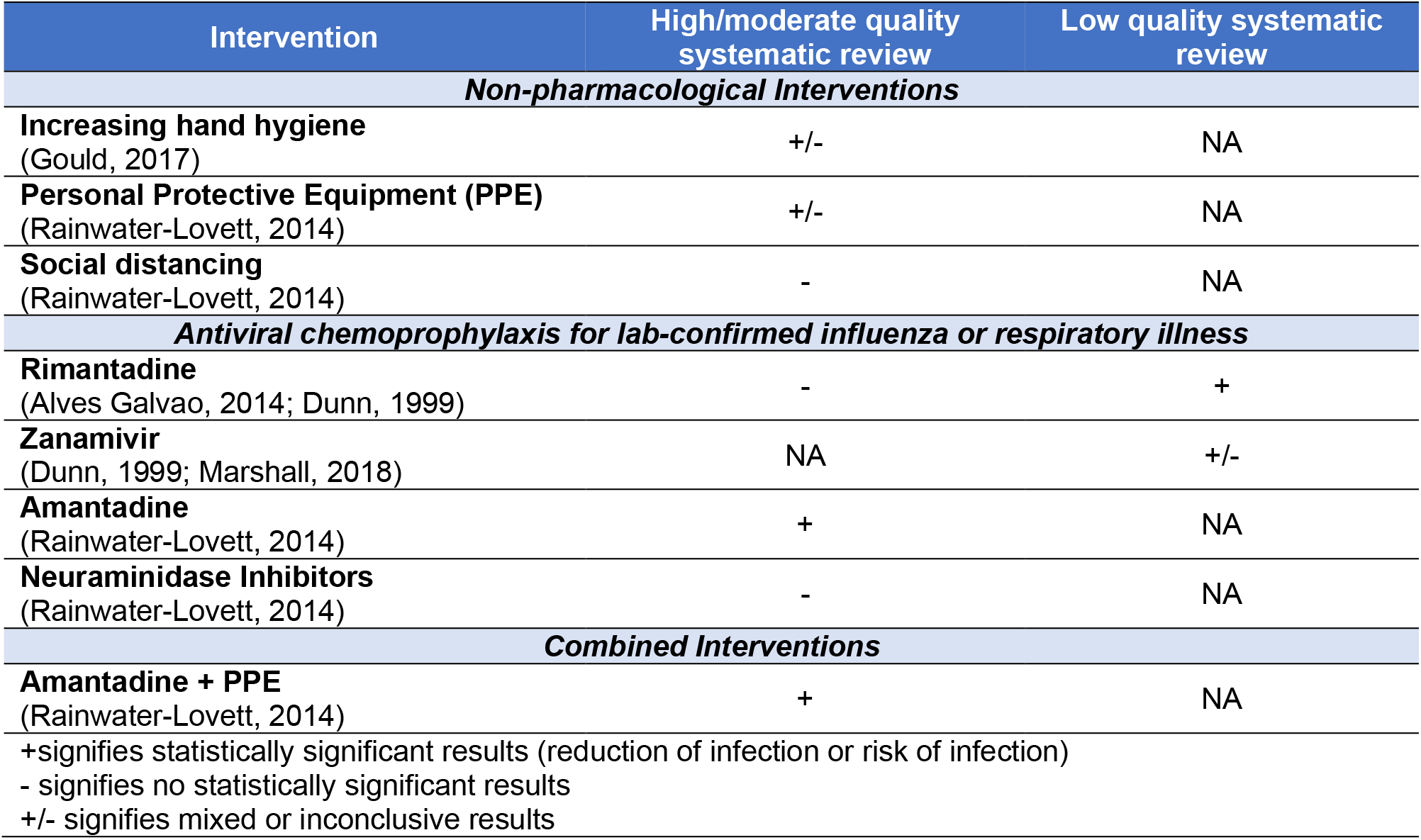
Summary of evidence for included systematic reviews.

## DISCUSSION

The WHO commissioned a rapid overview to address the urgent question of the infection prevention and control practices/measures for respiratory viruses in long-term care facilities that could be applied to COVID-19. A comprehensive literature search of both electronic databases and grey literature sources resulted in six included systematic reviews, none of which specifically focused on issues related to residents with respiratory illness and severe comorbidities or frailty. Furthermore, none focused on issues in LMIC or different cultural contexts.

Overall, the results of the included systematic reviews suggest that high quality evidence supports treating residents with antiviral chemoprophylaxis with adamantine, as well as adamantine in combination with personal protective equipment. For the rest of the strategies, there was either no evidence of effectiveness (e.g., social isolation) or mixed evidence of effectiveness (e.g., rimantadine, zanamivir, hand hygiene, personal protective equipment). The mixed evidence on hand hygiene and use of personal protective equipment does not imply these should not be used in outbreaks.

There are several limitations to the overview methods employed here, single screening and abstraction for example, however they were selected to thoughtfully tailor our methods according to our knowledge user needs and the urgent nature of the request to provide timely results.

## CONCLUSIONS

The current evidence suggests that with antiviral chemoprophylaxis with adamantine is effective in managing respiratory illness in residents of long-term care facilities. The rest of the strategies can be used in long-term care facilities, yet have limited evidence supporting their use.

## Data Availability

All datasets supporting the conclusions of this article are included within the article

## Acknowledgements

Jessie McGowan (literature search), Krystle Amog (report preparation), Chantal Williams (report preparation), Naveeta Ramkissoon (report preparation)

## Copyright claims/Disclaimers

The intellectual property rights in data and results generated from the work reported in this document are held in joint ownership between the Knowledge Translation Program team and the named Contributors.

Users are permitted to disseminate the data and results presented in this report provided that the dissemination (i) does not misrepresent the data, results, analyses or conclusions, and (ii) is consistent with academic practice, the rights of any third party publisher, and applicable laws. Any dissemination of the data and results from this document shall properly acknowledge the Knowledge Translation Program team and named Contributors.

## Funding Statement

This systematic review was commissioned and paid for by the World Health Organization and conducted through the Strategy for Patient Oriented-Research (SPOR) Evidence Alliance. The authors alone are responsible for the views expressed in this article and they do not necessarily represent the decisions, policy or views of the World Health Organization.

## APPENDIX 1 – PROTOCOL

**Team name:** Knowledge Translation Program of St. Michael’s Hospital, Unity Health Toronto (Drs. Tricco and Straus, contact)

**Query:** Preventing the transmission of coronavirus (COVID-19) in older adults aged 60 years and above living in long-term care

**Query Submitter:** Infection Prevention & Control, World Health Organization (WHO) Health Emergency (WHE) Programme

**Objective and research questions:**

The Infection Prevention & Control of the WHO WHE Programme has presented a query on the transmission of COVID-19 in older adults aged 60 years and above living in long-term care including privately paid for and publicly paid for settings with a 5-business day timeline. According to the World Health Organization, “long-term care covers those activities undertaken by others to ensure that people with, or at risk of, a significant ongoing loss of intrinsic capacity can maintain a level of functional ability consistent with their basic rights, fundamental freedoms and human dignity” (https://www.who.int/ageing/long-term-care/WHO-LTC-series-subsaharan-africa.pdf?ua=1) Examples of long-term care will include nursing homes, charitable homes, municipal homes, long-term care hospitals, long-term care facilities, skilled nursing facilities, convalescent homes, and assisted living facilities (https://www.canada.ca/en/health-canada/services/home-continuing-care/long-term-facilities-based-care.html).

The proposed research questions are:

What are the infection prevention and control practices/measures for preventing or reducing respiratory viruses (including coronavirus and influenza) in older adults aged 60 years and above living in long-term care?

How do infection prevention and control practices differ for adults aged 60 years and above living in long-term care with respiratory illness and severe comorbidities or frailty differ than those without such severe comorbidities/frailty?

How do infection prevention and control practices differ for adults aged 60 years and above living in long-term care with respiratory illness from low- and middle-income economy countries (LMIC) differ than those living in high-income economy countries and do differences exist across different cultural contexts?

### Research approach

The research question will be addressed through a rapid review informed by the methods proposed by the WHO Guide to rapid reviews (https://www.who.int/alliance-hpsr/resources/publications/rapid-review-guide/en/).

#### Protocol

Due to the urgent nature of the request and limited time frame to complete the work, this document will serve as the protocol for this query. If publication in a peer-reviewed journal is planned in the future, we will register this rapid review with the Open Science Framework (https://osf.io/).

#### Literature search

Comprehensive literature searches will be developed by an experienced librarian for MEDLINE, EMBASE, the Cochrane Library, biorxiv.org/medrxiv.org, and GIDEON (Global Infectious Diseases and Epidemiology Network). Grey (i.e., difficult to locate or unpublished) literature will be searched via clinicaltrials.gov. Due to the rapid timelines for this review a peer review of the literature search will not be conducted.

#### Eligibility criteria

The Eligibility criteria will follow the PICOST framework and will consist of:

<u>Population:</u> Individuals aged 60 years and above residing in long-term care facilities. The age cut-off for an older adult might be 50 years and above in some LMIC and/or cultural settings. As such, we will include these in level 1 screening of titles and abstracts and include anything deemed relevant at level 2 screening of full-text articles in an appendix.

<u>Interventions:</u> Any form of infection control and prevention, such as hand hygiene, respiratory hygiene/etiquette, personal protective equipment (for patients and health care providers), triage (on arrival), source control, isolation, daily monitoring/surveillance for signs and symptoms of respiratory illness (e.g., COVID-19) in residence, environmental cleaning, cleaning of linen and medical equipment used by patients, restrictions on resident movement and transportation, restrictions on visitors, restrictions on travel for health care providers and other long-term care facility staff, waste management, dead body management. Only those measures used to prevent and control respiratory illnesses, including influenza and coronavirus (e.g., COVID-19, MERS, SARS) will be included. Interventions focused on preventing bacterial respiratory outbreaks (e.g., strep, pneumonia, klebsiella) will be excluded.

<u>Comparator:</u> One of the interventions listed above or no intervention

<u>Outcomes:</u> Lab-confirmed respiratory illness due to the virus (e.g., SARS, MERS, COVID-19, influenza) [primary outcome], secondary bacterial infection, symptoms, secondary transmission (e.g., other patients, healthcare workers, visitors), goal concordant care, hospitalization, intensive-care unit (ICU) admission, mortality

<u>Study designs:</u> Due to the rapid nature of this request, we will limit inclusion to clinical practice guidelines and systematic reviews, using the Cochrane definition of a systematic review. If there is scant evidence from these study designs, we will expand inclusion to include the following study designs:

- Randomized controlled trials (RCTs)
- NRCTs (e.g., such as quasi-RCTs, non-randomized trials, interrupted time series, controlled before after),
- Observational studies (e.g., cohort, case control, cross-sectional)
- Case studies, case reports, and case series, including outbreak reports

<u>Time periods:</u> All periods of time and duration of follow-up will be included.

<u>Other limitations:</u> No other limitations will be imposed. If possible, we will translate studies written in languages other than English (e.g., Mandarin, Cantonese) that are deemed relevant.

#### Study selection process

In order to meet the requested timeline of 5 working days a streamlined approach to study selection will be employed. A screening form based on the eligibility criteria will be prepared and a brief calibration exercise will be conducted prior to citation and full-text screening. Screening will be completed by single reviewers using Synthesi.SR, the team’s proprietary online software (https://breakthroughkt.ca/login.php).

#### Data items and data abstraction process

Items for data abstraction will include study characteristics (e.g., duration of follow-up, study design, country of conduct, multi-center vs. single site, long-term care setting characteristics, such as availability of medical support, characteristics of care staff, family/community engagement, accommodation characteristics, collective practices), patient characteristics (e.g., mean age, age range, co-morbidities), intervention details (e.g., type of intervention, duration and frequency of intervention, timing of intervention), comparator details (e.g., comparator intervention, duration and frequency of intervention, timing of intervention), and outcome results (e.g., lab-confirmed viral respiratory infection, symptoms, secondary transmission, hospitalization, ICU admission, mortality) at the longest duration of follow-up. For the clinical practice guidelines, we will abstract the recommendations and level of evidence for reach recommendation.

Prior to data abstraction, we will complete a calibration exercise of the form amongst all reviewers using a random sample of 2 included articles. Following calibration, included studies will be abstracted by single reviewers.

#### Risk of bias appraisal

Risk of bias appraisal will be carried out by single reviewers using the AMSTAR-2 tool (https://amstar.ca/Amstar-2.php) for systematic reviews and the AGREE-2 tool (https://www.agreetrust.org/resource-centre/agree-reporting-checklist/) for clinical practice guidelines.

#### Synthesis

The synthesis will involve providing a descriptive summary of included studies with summary tables and detailed tables of study results. Tables of study results will be organized according to interventions of interest and reported outcomes and where available, information on relevant subgroups will be presented separately.

### Preliminary knowledge translation plan

The summary of results will be sent to the WHO and other relevant policy-makers as a brief summary report (1-5 pages) and 1-page policy brief (see http://www.cihr-irsc.gc.ca/e/documents/dsen-abstract-en.pdf for an example). We will work with the WHO team to consider submitting this paper to an open-access, peer-reviewed journal for publication (e.g. British Medical Journal).

### Timeline (from the point of official approval)

Five business days (March 16, 2020)

### Updates provided to the WHO

Daily emails will be sent to the WHO

### Funding

Funding will be obtained from the WHO and the Canadian Institutes of Health Research Strategy for Patient Oriented Research Evidence Alliance (https://sporevidencealliance.ca/).

## APPENDIX 2 – MEDLINE SEARCH STRATEGY

1. respiratory tract infections/ or exp bronchitis/ or exp common cold/ or exp influenza, human/ or laryngitis/ or exp pharyngitis/ or exp pleurisy/ or exp pneumonia/ or exp rhinitis/ or exp rhinoscleroma/ or exp severe acute respiratory syndrome/ or exp sinusitis/ or exp supraglottitis/ or exp tracheitis/ or exp whooping cough/
2. coronaviridae infections/ or coronavirus infections/ or SARS Virus/
3. (coronavirus* or “corona virus*” or mers or “middle east respiratory syndrome*” or “Severe Acute Respiratory Syndrome*” or SARS or CoV or SARS-CoV or MERS-CoV or 2019-nCoV or COVID-19 or “2019 novel coronavirus disease” or “2019 ncov disease” or “2019 ncov infection” or “coronavirus disease 19” or “severe acute respiratory syndrome coronavirus 2” or “severe acute respiratory syndrome coronavirus 2” or “wuhan” or “sars cov 2”).tw,kf.
4. (flu or influenza or “respiratory tract infection*” or “respiratory infection*” or bronchitis or “common cold” or laryngitis or pharyngitis or pneumonia or rhinitis or rhinoscleroma or sinusitis or supraglottitis or tracheitis or “whooping cough”).tw,kf.
5. or/1-4
6. pc.fs.
7. exp Infection Control/ or secondary prevention/
8. exp hand hygiene/ or hygiene/
9. (prevent* or “respiratory hygiene” or “respiratory etiquette “ or “cough etiquette” or “Hand Hygiene” or “hand wash*” or “handwash*” or “patient isolation” or “quarant*” or “infection control” or “blood safety” or steril* or disinfect* or “contract tracing” or “disease notification” or fumigat* or “personal protective equipment” or triage or “source control” or isolation or “daily monitoring” or surveillance or “waste management” or cadaver or body or corpse or “face mask*” or facemask* or “social distanc*” or housekeeping). tw,kw.
10. (clean* adj3 (linen or equipment or environment)).tw,kf.
11. (restrict* adj3 (resident* or patient* or visit* or family or travel* or staff or provider* or employee*)).tw,kf.
12. personal protective equipment/
13. Housekeeping, Hospital/
14. Waste Management/
15. patient isolation/
16. triage/
17. Cadaver/
18. or/6-17
19. 5 and 18
20. Long-Term Care/ or exp Nursing Homes/ or Homes for the Aged/ or Assisted Living Facilities/
21. (“long-term care” or “long term care” or “senior* home*” or “senior* residence*” or “nursing home*” or “old age home*” or “old age residence*” or “home* for the aged”).tw,kf. (48416)
22. 20 or 21
23. 19 and 22

## APPENDIX 3 – SYSTEMATIC REVIEW CHARACTERISTICS

| Author, Year<br>Country | Methods | Included Studies<br>and Population | Setting | Intervention |
| --- | --- | --- | --- | --- |
| Alves Galvao,<br>2014<br>Brazil | Randomised controlled trials (RCTs) and quasi-RCTs comparing amantadine or rimantadine, or both, with placebo, control drugs, different doses or schedules of amantadine or rimantadine, or both, or no intervention, in children and the elderly. We included studies where at least 75% of the population was up to 19 years of age, or 65 years of age or older. We also included trials with a wider age range where data by age subgroups were available. Comparisons of amantadine or rimantadine, or both, to placebo, control drugs, other antivirals, no interventions or different doses of amantadine or rimantadine, or both, as prophylaxis and/or treatment for influenza A. | 3 studies<br>N = 908 | Nursing home | Trials assessing Amantadine and/or rimantadine for preventing influenza |
| Gould, 2017<br>Canada | We considered randomised trials (RCTs), non-randomised trials, controlled before-after studies (CBAs) and interrupted time series (ITS) studies. To be eligible for review, ITS studies had to demonstrate a clearly defined point in time when the intervention occurred, and include at least three data collection points both before and after the intervention. We considered studies where the participants or target groups were nurses, doctors and other healthcare workers in any hospital, nursing home, long-term care facility or community healthcare setting in any country | 26 studies<br>N = NR | 2 studies conducted in long-term care facilities, 1 in a primary care setting, and the remaining 23 studies were conducted in acute care hospitals on general wards and/or critical care units | Multimodal campaigns featuring complex interventions; alcohol based hand rub; performance based feedback; education; cues |
| **Marshall, 2018**<br>**NR** | Literature search was conducted in PubMed, Embase and Cochrane and Centers for Disease Control and Prevention (CDC), Association for Professionals in Infection Control (APIC) and Centers for Medicare & Medicaid Services (CMS) websites. To provide current guidelines for LTC influenza outbreak management, focusing on the pharmacist's role in management, particularly regarding oseltamivir use and potential adverse drug reactions (ADRs). | NR<br>N = NR | Long-term care facilities | CDC, APIC and CMS recommend antiviral treatment for residents with confirmed or suspected influenza and antiviral chemoprophylaxis for non-ill residents exposed during outbreak. |
| **Dunn, 1999**<br>**New Zealand** | Medical literature published in any language since 1966 on zanamivir, identified using AdisBase (a proprietary database of AdisInternational, Auckland, New Zealand), Medline and EMBASE. | 1 study<br>N = 140 | Nursing home | Prophylactic 14-day regimens of zanamivir 10mg by inhalation plus 3.2mg intranasally twice daily or rimantadine 100mg orally once daily (influenza A); zanamivir only for influenza B. |
|  | Additional references were identified from the reference lists of published articles. Bibliographical information, including contributory unpublished data, was also requested from the company developing the drug. Studies in patients with influenza A or influenza B who received zanamivir, and studies in which the drug was given prophylactically. Inclusion of studies was based mainly on the methods section of the trials. When available, large, well-controlled trials with appropriate statistical methodology were preferred. Relevant pharmacodynamic and pharmacokinetic data are also included. |  |  |  |
| Volkman, 2012<br>USA | A literature review was conducted in September of 2010 using the Cumulative Index to Nursing and Allied Health Literature (CINAHL), Healthstar, PubMed, and MEDLINE databases for years 1966 through 2010, as well as an Internet search for existing state guidelines, national recommendations, and accreditation standards. The search terms used were as | 33 studies, N = NR | Long-term care facilities (LTCFs) - defined as residential institutions that provide care to people who are unable to live independently. There are various types/classifications of LTCFs, based on the patient population and services provided. Examples include nursing homes, skilled nursing facilities, long-term acute care | Infection prevention program in place for day-to-day activities as well as during disasters; having coverage by a designated infection preventionist (IP) (LTCFs employ a certified or trained IP); LTCFs must develop and maintain an emergency operations plan that addresses all hazards, including biologic threats. The facility EOP needs to be coordinated with local, state, and federal plans. LTCFs must assess their disaster |
|  | follows: long-term care, nursing home, disaster preparedness, infection prevention, emergency preparedness, hurricane, pandemic influenza, and stockpiling. Only English language articles or documents from peer-reviewed journals, national organizations, or accrediting agencies were included. In addition, the snowballing technique was used to identify sources. |  | facilities, psychiatric institutions, foster and group homes, retirement homes, and rehabilitation centers. The majority of LTCFs are nursing homes that house elderly and chronically ill individuals at high risk for infection. Therefore, this document will focus primarily on nursing homes as LTCFs. However, infection prevention recommendations in this article may be modified and applied for other LTCFs. | readiness, Each LTCF should designate a person whose responsibility it is to create, coordinate, and track staff training on emergency management, Ideally, LTCF should have the ability to be self-sustaining for at least 96 hours. To do this, the facility EOP must address resource assessment and management. |
| **Rainwater-Lovett, 2013**<br>**USA** | PubMed was searched. Studies of any design reporting influenza outbreaks among elderly individuals in LTCFs were considered for inclusion. During full text review, articles were included if they contained the number of influenza cases occurring on a specific date or within a 3-day period, such as through an epidemic curve or a line list of symptom onset dates | 37 studies<br>N = NR | Long term care facilities - defined as any residential environment that housed older adults or elderly individuals with the assistance of medical staff and included facilities referred to as 'assisted living' or 'nursing homes' but excluded community centers and daytime-only facilities serving older adults living in the outside community. | Chemoprophylaxis was defined as offering antiviral drugs to asymptomatic individuals in the facility; Personal protective equipment (PPE) included glove and mask use, hand hygiene, and droplet precautions; Social distancing included patient isolation and restrictions on staff, visitors, admissions and ward transfers. Isolation was defined as restriction of movement within the facility by symptomatic individuals through the use of room, unit, or ward quarantine or cohorting. |

## APPENDIX 4 – QUALITY APPRAISAL RESULTS FOR SYSTEMATIC REVIEWS AND CLINICAL PRACTICE GUIDELINES

| Author, Year | 1. PICO components | 2. A priori design | 3. Rationale for study selection | 4. Literature search | 5. Duplicate Selection | 6. Duplicate Abstraction | 7. List of excluded studies | 8. Description of included studies | 9a. Risk of Bias assessment in RCTs | 9b. Risk of bias assessment in non-randomized studies | 10. Funding sources | 11. Appropriate MA methods | 12. Used RoB in MA | 13. Used RoB in interpreting results | 14. Discussion of heterogeneity | 15. Publication bias | 16. Conflict of Interest | Overall Rating |
| --- | --- | --- | --- | --- | --- | --- | --- | --- | --- | --- | --- | --- | --- | --- | --- | --- | --- | --- |
| **Alves Galvao, 2014** | Yes | Yes | Yes | Yes | Yes | Yes | Yes | Yes | Yes | Yes | Yes | Yes | Yes | Yes | Yes | Yes | Yes | High |
| **Dunn, 1999** | No | No | Yes | Yes | No | No | No | Yes | No | No | Yes | NA | NA | No | NA | NA | Yes | Low |
| **Gould, 2017** | Yes | Yes | Yes | Yes | Yes | Yes | Yes | Yes | Yes | Yes | Yes | NA | NA | Yes | NA | NA | Yes | High |
| **Marshall, 2018** | Yes | No | No | Yes | No | No | No | Partial | No | No | Yes | NA | NA | No | NA | NA | No | Low |
|  |  |  |  |  |  |  |  | Yes |  |  |  |  |  |  |  |  |  |  |
| **Rainwater-Lovett, 2014** | Yes | Partial<br>Yes | Yes | Yes | Yes | Yes | No | Yes | No | No | Yes | Yes | No | No | No | No | Yes | Moderate |
| **Volkman, 2012** | Yes | No | Yes | Yes | No | No | No | Yes | No | No | No | NA | NA | No | NA | NA | Yes | Low |

NA – 'Not Applicable'; no meta-analysis was conducted in these reviews

## APPENDIX 5 – SYSTEMATIC REVIEW RESULTS

| Outcome specific results | Overall Conclusions |
| --- | --- |
| **Alves Galvao, 2014; AMSTAR Rating: High, Condition: Influenza** |  |
| No protective effect of rimantadine was seen regarding the prophylaxis of influenza in the elderly: 191 participants, RR 0.74; 95% CI 0.13 to 4.07; no protective effect of rimantadine prophylaxis in the occurrence of cases of influenza persisted (103 participants, RR 0.45; 95% CI 0.14 to 1.41); A reduced rimantadine dose of 100 mg/day was comparable to the full dose of 200 mg daily for prophylaxis of influenza in the elderly, although a wide CI was verified (54 participants, RR 0.93; 95% CI 0.21 to 4.20) | Rimantadine did not show any prophylactic effect in the elderly. The quality of evidence was very low: 103 participants (RR 0.45; 95% CI 0.14 to 1.41). The assumed risk was 17 per 100. The corresponding risk in the rimantadine group was 7 per 100 (95% CI 2 to 23). There was no evidence of adverse effects caused by treatment with amantadine or rimantadine |
| **Dunn, 1999; AMSTAR Rating: Low, Condition: Influenza A/B** |  |
| During the 14-day prophylaxis period, there were 9 new cases of respiratory illness in volunteers in units affected by influenza A. None of these were confirmed as influenza in 65 volunteers receiving zanamivir, whereas there was 1 confirmed case among the 23 volunteers receiving rimantadine. Of the 35 volunteers randomised to zanamivir and the 17 randomised to supportive care on units affected by influenza B, there was 1 case of laboratory-confirmed influenza in the supportive care group. | None reported |
| **Gould, 2017; AMSTAR Rating: High, Condition: Viral respiratory illness** |  |
| 1 RCT reported reduced respiratory outbreaks and MRSA infections requiring hospitalisation (IRR 0.12 to 0.61) favouring the intervention, while 1 ITS study reported no reduction in MRSA clinical isolates or infection. 1 RCT reported reductions of 0.27 to 0.77 cases per 1000 resident-days in serious infections, pneumonia and death in the intervention group compared to no change or an increase of 0.57 cases per 1000 resident-days in the control group | With the identified variability in certainty of evidence, interventions, and methods, there remains an urgent need to undertake methodologically robust research to explore the effectiveness of multimodal versus simpler interventions to increase hand hygiene compliance, and to identify which components of multimodal interventions or combinations of strategies are most effective in a particular context |
| **Marshall, 2018; AMSTAR Rating: Low, Condition: Influenza** |  |
| The CDC, APIC and CMS recommend antiviral treatment for residents with confirmed or suspected influenza and antiviral | The LTC pharmacist can play a key role in influenza management by helping identify outbreaks, distributing |
| chemoprophylaxis for non-ill residents exposed during outbreaks.<br>Oseltamivir, zanamivir and peramivir were the antivirals recommended for the 2017-2018 season. Oseltamivir has been the prescribed antiviral in LTC despite mixed reports of effectiveness in residents. Treatment should begin within 48 hours of symptom onset or exposure. Oseltamivir dosage should be decreased for patients with creatinine clearance less than 60 mL/minute and it's not recommended for end stage renal disease patients not on dialysis. The most common oseltamivir prophylaxis/treatment ADRs reported were nausea, vomiting, headache and pain but neuropsychiatric ADRs of hallucinations, abnormal behavior, confused state, and delirium have occurred, resulting in discontinuation in some cases. Of 10,218 cumulative ADRs reported to the FDA, 2,412 were psychiatric and 2,019 were nervous system; 12% of the cumulative reports involved patients aged 65 and older | medication in a timely manner, providing information regarding oseltamivir benefits/risks and dosages, identifying potential ADRs, and suggesting discontinuation when warranted |
| **Rainwater-Lovett, 2014; AMSTAR Rating: High, Condition: Influenza** |  |
| Individuals in facilities where chemoprophylaxis was used were significantly less likely to develop influenza A or B than those in facilities with no interventions [odds ratio (OR) 0.48, 95% CI: 0.28, 0.84]. Considered by drug class, amantadine significantly reduced infection risk (OR 0.22, 95% CI: 0.12, 0.42), while neuraminidase inhibitors did not show a significant effect. Although NPI showed no significant effect, the results suggest that personal protective equipment may produce modest protective effects. After adjusting for NPI, use of any antiviral drug halved attack rates among outbreaks consisting of influenza A cases [OR: 0.52 (95% CI: 0.29, 0.93)] and influenza A and B cases [OR: 0.48 (95% CI: 0.28, 0.84)] compared with outbreaks that did not implement influenza control measures. More specifically, the use of amantadine produced statistically significant protective effects among outbreaks with cases of influenza A [OR: 0.33 (95% CI: 0.17, 0.62)] and influenza | This analysis provides further evidence that antiviral prophylaxis is one of the most effective ways to control influenza in populations at high risk of complications and where vaccine efficacy is reduced. While antiviral prophylaxis is highly effective, it may fail in the face of a novel, resistant strain. In such a scenario, NPI will be our only option for control and the importance of understanding which measures are most effective, and how effective they are, is paramount. Antiviral prophylaxis significantly reduced influenza attack rates, reducing the odds of developing influenza by 50% among LTCF residents. While our results were consistent with a protective effect of PPE, this effect was not statistically significant. We suspect the lack of statistically significant protective effects of PPE and social distancing was the result of broad definitions as NPI were rarely reported in detail |
| A and B [OR: 0.27 (95% CI: 0.14, 0.48)]. Combined use of amantadine and neuraminidase inhibitors consistently demonstrated protective effects, although not statistically significant, but surprisingly neuraminidase inhibitors alone did not. Social distancing (influenza A: OR 1.05, 95% CI: 0.53, 2.16; influenza A and B: OR 1.07, 95% CI: 0.58, 1.90) and PPE (influenza A: OR 0.75, 95% CI: 0.33, 1.61; influenza A and B: OR 0.99, 95% CI: 0.49, 1.93) were not associated with significant changes in influenza attack rates. |  |
| **Volkman, 2012; AMSTAR Rating:** Low, **Condition:** Viral infections |  |
| None applicable | Whereas there were multiple publications related to the difficulties and risk factors LTCFs face in disasters, there were no publications that specifically addressed infection prevention in disasters or planning specific to infection prevention concerns in disasters in long-term care. All health care settings need to have comprehensive emergency operations plans that address all hazards. LTCFs have unique challenges not experienced by other health care settings, and these issues need to be addressed as part of facility emergency management planning. Protocols need to be in place that will minimize infection transmission risk among patients, visitors, and staff during disasters. This article summarizes disaster planning considerations related to infection prevention for LTCFs, including having an infection prevention program, developing and assessing an emergency operations plan that is coordinated with regional and federal plans, creating and implementing an infection prevention education program related to disaster preparedness, and managing supplies. LTCFs should use this article to develop and assess their EOP and staff training as it relates to infection prevention procedures and protocols. |

## Notes

### Competing Interest Statement

The authors have declared no competing interest.

